# Device-led versus human-led feedback on chest compressions for cardiopulmonary resuscitation and providers’ experience and preference: a randomised crossover study

**DOI:** 10.1101/2022.09.25.22280283

**Authors:** Muhaimin Noor Azhar, Aida Bustam, Khadijah Poh, Keng Sheng Chew, Asraff Azman, Anhar Kamarudin, Aliyah Zambri

**Affiliations:** Academic Unit of Emergency Medicine, Faculty of Medicine, University of Malaya, Kuala Lumpur, Malaysia; Faculty of Medicine and Health Sciences, Universiti Malaysia Sarawak Kota Samarahan, Sarawak, Malaysia; Department of Emergency Medicine, University Malaya Medical Center, Kuala Lumpur, Malaysia

## Abstract

**Background:** High cardiopulmonary resuscitation (CPR) quality is associated with better patient survival from cardiac arrest. However, CPR providers may not have an accurate perception of the depth and rate of their chest compressions (CC). Realtime feedback during resuscitation improves CPR quality compared to no feedback. Evidence comparing audio-visual feedback device (AVF) and team leader’s feedback (TLF) in improving CPR performance is limited and conflicting.

**Methodology:** We performed a randomized crossover study to evaluate CC performance with AVF and TLF. Seventy participants performed CC for 1 minute on a CPR manikin connected to ZOLL R series defibrillator with CPR-sensing capability in a randomised crossover sequence. We interviewed participants to explore their perception and preference with both feedback methods.

**Results:** Mean CC rate was higher with AVF than with TLF (121.8 min^-1^ ± 17.7 vs. 117.4 min^-1^ ± 13.5, *p* = 0.005). There was no significant difference in proportions of participants performing CC within the recommended rate of 100-120 beats per minute between AVF and TLF (48.6% and 51.4%, *p* = 0.824). Overall, CC depth was below the recommended target regardless of feedback method with mean CC depth of 4.4 cm ± 0.8 in AVF and 4.3 cm ± 0.9 in TLF respectively (*p* = 0.479). Most participants felt that TLF was easier to follow, more motivating and preferable compared to AVF. Those who preferred TLF performed CC at rates above the recommended range with AVF compared to TLF (124.1 min^-1^ ± 19.4 versus 118.2 min^-1^ ± 14.9, *p* = 0.004).

**Conclusion:** A well-trained team leader is as effective as an AVF device in leading high-quality CC. CPR providers’ performance may be influenced by their preferred feedback method.

## Introduction

Cardiac arrest is one of the major public health issues worldwide with a global incidence of as high as 110.8 cases per 100,000 people (1). One of the life-saving interventions in the management of cardiac arrest is cardiopulmonary resuscitation (CPR). CPR involves the provision of chest compressions and ventilations to deliver oxygenated blood to the brain. However, patient survival rates from cardiac arrest remain low even among those who received CPR (1). The quality of CPR provided during resuscitation has an important association with patient survival from cardiac arrest (2-4). CPR guidelines have emphasised on the delivery of high-quality CPR which includes targeted optimal compression rate, adequate compression depth and minimising interruptions between chest compression (5). The 2015 American Heart Association (AHA) and European Resuscitation Council (ERC) CPR guidelines recommend an optimum chest compression rate of 100 to 120 min^-1^ and a chest compression depth of 5 to 6 cm (6, 7).

Despite an increasing knowledge about CPR physiology in optimising blood flow during cardiac arrest (8), and of its impact on neurological and survival outcome of patients with cardiac arrest (9), challenges still exist in its implementation. It has been shown that high quality CPR is infrequently delivered in clinical practice even among well-trained healthcare providers (10, 11). CPR providers may not have an accurate perception of the depth and rate of their chest compressions (12). This reinforces the need to monitor and improve the quality of CPR during resuscitation of cardiac arrest patients. Real-time monitoring and prompt feedback on CPR quality during a resuscitative effort can guide real-time corrective measures by the CPR provider. Traditionally, this is done through subjective visual assessment by a team leader or another rescuer who monitors and provides guidance and feedback on the provider’s CPR performance. Team leaders in the context of resuscitation is considered a role adopted by one member of the team who assumes responsibility for managing a cardiac arrest. Recently, CPR feedback devices equipped with sensors and accelerometer technology have become available that enable the detection of CPR metrics such as rate and depth of chest compressions. These devices provide automated audio feedback, and/or visual feedback in the form of graphs and numbers, to alert the provider when the CPR metric values fall outside of the pre-programmed range.

Previous studies investigating the effectiveness of CPR feedback devices have found improved CPR quality when compared with no feedbacks (13-18). CPR feedback devices can be useful as part of a strategy to improve CPR quality during resuscitation. However, well-trained CPR providers may remain as the predominant and effective team leaders during CPR, particularly in limited-resource environments (19). Evidence for CPR performance comparing feedback device with human-led feedback is still limited, with one study reporting improved CPR quality with feedback device (20), and another study that otherwise found comparable CPR quality between device-led and human-led feedback (21). Furthermore, CPR providers’ comparative perception of experience and preference with both methods of feedback during CPR have yet to be explored.

In this study, we aimed to (1) determine whether the use of an audio-visual CPR feedback device compared with team leader feedback improves chest compression quality in a simulation setting, (2) explore CPR providers’ perception and preference with both feedback methods, and (3) evaluate if providers’ perception or preference is associated with quality of chest compressions.

## Materials and Methods

### Study design and setting

This was a manikin-based simulation study comparing chest compression (CC) performance by participants with audio-visual feedback (AVF) versus team leader’s feedback (TLF). We performed a randomized 2-sequence, 2-intervention periods crossover study design. The study flow is shown in Fig. 1. This study was conducted from November to December 2019 in an emergency department (ED) of a university-affiliated hospital in Malaysia. A crossover study design was chosen because of advantages over a parallel-group randomized controlled design: each participant could serve as his or her own matched control and therefore reducing within-participant variation, a smaller sample size is required to detect meaningful effect at the same level of statistical power as a parallel design, and participants in this crossover study could express their preferences by comparing their experiences of the two interventions (22). This study is reported according to the Consolidated Standards of Reporting Trials approach with the extension for simulation-based research. This study was granted ethics approval from the hospital Medical Research Ethics Committee (MREC 201919-6992) and the Malaysian National Medical Research Register (NMRR-19-1174-48443) in accordance with the International Conference on Harmonization - Guidelines for Good Clinical Practice (ICH-GCP) and Declaration of Helsinki.

**Fig 1.** Study flowchart TLF: team-leader feedback, AVF: audio-visual feedback, CC: chest compression

### Participant selection and recruitment

Eligible participants were Basic Life Support (BLS)-certified ED healthcare personnel and students. Exclusion criteria were previous experience with using AVF devices and any physical disabilities, injuries and/ or chronic medical illnesses that may impair the ability to perform high quality CC. Due to work scheduling constraints, participant recruitment was by convenience sampling. All recruited participants enrolled voluntarily and signed an informed consent form.

### Randomization and crossover

A study investigator not involved in participant recruitment and study interventions, randomized the participants using computer-generated block-stratified random sequence and concealed the lists in opaque envelops. The stratification was by profession using block size of 4. Enrolled participants were randomized in a 1:1 allocation ratio into AVF-led CC followed by TLF-led CC (Group 1), or TLF-led CC followed by AVF-led CC (Group 2). We allocated a 10-minute washout interval between intervention periods to minimize participant fatigue.

### Equipment

The CPR manikin used was Little Anne™ (Laerdal, Orpington, UK) with a spring constant of 4.46 kg/cm and a 22.3 kg-force required to press 5 cm and maximum compression depth of 7 cm. The CPR manikin was connected to a ZOLL R Series Plus® (ZOLL Medical Corporation) defibrillator with CPR-sensing capability and AVF aptitude algorithm based on the 2015 AHA guidelines. The CPR Dashboard™ using Real CPR Help® technology in the device provided real-time AVF on rate, depth, and release of each compression via an accelerometer.

### Interventions

The CPR manikin was placed on the floor to avert mattress compressibility as a confounding variable. Participants were instructed to perform CC as they would usually perform in real practice, but with a small CPR sensor placed underneath their hands on the manikin’s chest. AVF configuration was enabled during the AVF-led intervention and was disabled during the TLF-led intervention. For the TLF-led intervention, three certified BLS trainers were assigned as team leaders. Based on their availability, one of the three team leaders conducted the TLF-led session and provided verbal feedback for compression rate and depth. The team leader was not present during the AVF-led intervention to minimize bias. Each participant performed a 1-minute chest compression in each intervention period.

### Data collection and outcome measures

We collected participants’ demographic data comprising of age, gender, and profession. The primary outcome measured were CC rate (min^-1^) and depth (cm). We extracted the CC rate and depth data stored in the built-in memory storage of the defibrillator using the ZOLL CodeNet software provided by the manufacturer. For secondary outcome measures, we interviewed each participant immediately after completion of both interventions to obtain qualitative feedback regarding their perception and experience. Each interview was audio-recorded and later transcribed for analysis to look for common themes. We also asked if they felt they performed better with AVF or with TLF, and their preferred method of feedback during resuscitation.

### Sample size calculation

An earlier 2 × 2 crossover study of CC with and without feedback device showed mean chest compression rates of 113 min^-1^ (± 7) and 113 min^-1^ (± 13) respectively (17). Based on this and the AHA-recommended CC target rate range of 100 to 120 min^-1^, we defined the minimal clinically significant difference in CC rate as 7 min^-1^ for this study (since difference of >7 min^-1^ would exceed the recommended CC target rate). Using sample size calculation for two-intervention crossover study, a total of 57 participants will have 80% power to detect a difference in mean of 7 min^-1^ at a two-sided 0.05 significance level, if the within-participant standard deviation is 13 min^-1^ (for a more conservative estimate). Sample size calculation was performed with a web-based sample size calculator software (23). We recruited 70 participants to include an estimated 20% dropout rate.

### Statistical analysis

Statistical analysis was performed with SPSS version 26 for Mac OS. Demographic characteristics were analyzed with descriptive statistics. Test of normality was performed by examining skewness z-score, kurtosis z-score, and Shapiro-Wilk test. Parametric data was analyzed with paired t-test and non-parametric data with Wilcoxon signed rank test. Categorical data was analyzed with McNemar’s test. All tests were performed at the significance level of *p* < 0.05.

## Results

### Participants

A total of 70 participants (mean age 29.17 ± 5.36 years) were enrolled and no participants were excluded from the study. Thirty-nine (55.7%) participants were female. The participants were made up of 31 doctors (44.3%), 16 nurses (22.9%), 9 paramedics (12.9%) and 14 medical students (20%).

### Chest compression performance

The comparison of CC rate and depth between TLF-led CC and AVF-led CC is shown in **Table 1**. The mean CC rate was higher in AVF-led CC compared with TLF-led CC (121.75 ± 17.66 min^-1^ vs. 117.43 ± 13.45 min^-1^, *p* = 0.005). The mean CC rate with AVF was also slightly above the recommended target rate (>120 min^-1^). There was no significant difference between AVF-led CC and TLF-led CC in the proportion of participants who performed CPR at rates within and not within the recommended target rate (*p* = 0.824). Mean CC depth performed by participants did not meet the recommended target of 5-6 cm in both interventions (4.37 ± 0.78 cm in AVF-led CC and 4.33 ± 0.92 cm in TLF-led CC, *p* = 0.479). There was no significant difference between AVF-led CC and TLF-led CC in the proportion of participants who performed CPR with depth within and not within the recommended target (*p* = 0.754).

**Table 1.** Comparison of chest compression rate and depth between TLF-led CC and AVF-led CC, and proportion of participants performing below, within or above recommended targets in guideline.

| CC variables | AVF-led | TLF-led | p-value |
| --- | --- | --- | --- |
| **Rate, min<sup>-1</sup>** |  |  |  |
| Mean (SD) | 121.8 (17.7) | 117.4 (13.5) | 0.005<sup>a</sup> |
| 95% CI | 117.5, 126.0 | 114.2, 120.6 |  |
| **CC rate performance categories, n (%)** |  |  |  |
| Below target (<100) | 4 (5.7) | 5 (7.1) |  |
| Within target (100-120) | 34 (48.6) | 36 (51.4) | 0.824<sup>b</sup> |
| Above target (>120) | 32 (45.7) | 29 (41.4) |  |
| **Depth, cm** |  |  |  |
| Mean (SD) | 4.4 (0.8) | 4.3 (0.9) | 0.479<sup>a</sup> |
| 95% CI | 4.2, 4.6 | 4.1, 4.6 |  |
| **CC depth performance categories, n (%)** |  |  |  |
| Below target (<5) | 52 (74.3) | 49 (70.0) |  |
| Within target (5-6) | 18 (25.7) | 20 (28.6) | 0.754<sup>b</sup> |
| Above target (>6) | 0 (0.0) | 1 (1.4) |  |

<sup>a</sup>Paired t-test.
<sup>b</sup>McNemar's test performed to evaluate the difference in proportion with CC variables categorized into within target and not within target.
TLF: team-leader feedback, AVF: audio-visual feedback, CC: chest compression, SD: standard deviation, CI: confidence interval

### Exploration of participants’ feedback

Three common themes emerged from the subjective participant feedback: focus, comprehension, and motivation (**Table 2**).

**Table 2.**
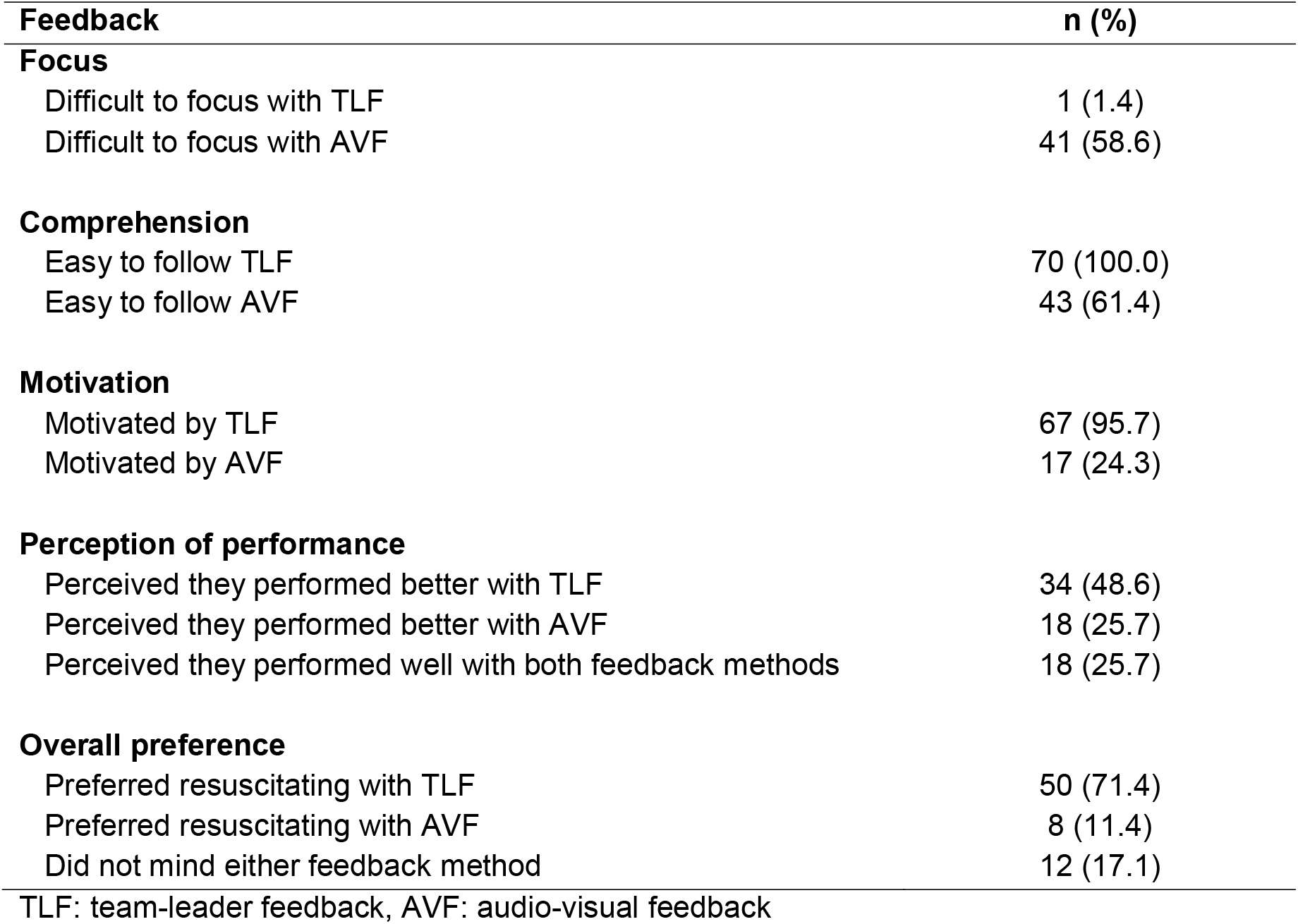
Participants’ feedback (N = 70)

| Feedback | n (%) |
| --- | --- |
| **Focus** |  |
| Difficult to focus with TLF | 1 (1.4) |
| Difficult to focus with AVF | 41 (58.6) |
| **Comprehension** |  |
| Easy to follow TLF | 70 (100.0) |
| Easy to follow AVF | 43 (61.4) |
| **Motivation** |  |
| Motivated by TLF | 67 (95.7) |
| Motivated by AVF | 17 (24.3) |
| **Perception of performance** |  |
| Perceived they performed better with TLF | 34 (48.6) |
| Perceived they performed better with AVF | 18 (25.7) |
| Perceived they performed well with both feedback methods | 18 (25.7) |
| **Overall preference** |  |
| Preferred resuscitating with TLF | 50 (71.4) |
| Preferred resuscitating with AVF | 8 (11.4) |
| Did not mind either feedback method | 12 (17.1) |

TLF: team-leader feedback, AVF: audio-visual feedback

### Focus

Forty-one participants (58.60%) reported difficulty to concurrently focus on delivering high quality CC and AVF as illustrated below:

> *“With the machine, I needed to keep counting the rate in my head, while looking at the monitor. I needed to focus on many things at once. Whereas with the team leader, I could concentrate better by just listening to one voice*.*”*
>
> *“Better with the team leader. With the machine, I needed to look at the monitor, listen and follow the voice prompts. I cannot perform many things simultaneously – to see, to listen, to interpret all at the same time*.*”*

### Comprehension

All participants reported that it was easy to follow TLF instructions as opposed to only 43 (61.4%) who found AVF easily understood. Some participants were inclined to push faster when the AVF prompted them to “push harder”. For example:

> *“When the machine says push harder, I have the tendency to push faster just to fulfil the aim set by the machine*.*”*
>
> *“I prefer team leader even though the machine seems more objective, but the team leader’s instruction is simpler and easier to understand. The machine can be distracting*.*”*

### Motivation

Sixty-seven participants (95.7%) felt motivated during TLF-led CC compared to 17 (24.3%) participants in AVF-led CC. Examples include:

> *“When I was performing CPR with the machine, when the machine said “push harder” I panicked and had the tendency to push harder. Whereas, with team leader, he would encourage me by saying “good compressions”, and so, it made me calmer in doing CPR*.*”*
>
> *“I prefer the team leader because she gives constructive feedback. The team leader’s voice and commands are clear and it is not dull or monotonous voice like that of the machine. When we perform chest compressions, we have to be very energetic. A human voice will keep us motivated and energetic to give our best during CC*.*”*

### Perception of performance

Thirty-four (48.6%) participants felt they performed CC better with TLF, whereas 18 (25.7%) participants felt they performed better with AVF, and 18 (25.7%) participants felt they performed well with both feedback methods. For instance:

> “*I think I did better chest compressions guided by team leader, as I can solely focus only on my compressions without having to multitask*.*”*
>
> *“I think my compressions were more effective with the machine. It gave me objective feedback if I need to push harder or faster. I feel my compressions are much better because of that*.*”*

### Overall preference

Overall, 50 (71.4%) participants preferred instructions from TLF compared to only 8 (11.4%) participants who preferred instructions from AVF. Reasons for preferring TLF included the provision of a more comprehensive and holistic feedback. For example:

> *“I prefer the team leader because he gave me very specific and personalized feedback like I needed to straighten and lock my elbows. Whereas for the machine, it only tells me about the depth and rate of compression”*

Those who preferred AVF generally cited “more objective and straightforward feedback” as the reason.

> *“I prefer the instructions given by the machine. Because, first, I can observe how much I need to improve – the rate, the depth, I know exactly how much I need to improve*.
>
> *Whereas with team leader, while it is also helpful, when he or she says push harder, but I am not seeing how much exactly I need to improve on*.*”*

### Participants’ perceived performance and preference, and chest compression quality

Participants who perceived that they performed better with TLF, and those who preferred TLF, were noted to perform CC at a mean rate significantly higher and above the recommended target rate when performing with AVF (**Table 3**). Participants who perceived that they performed better with AVF, and those who preferred AVF, showed similar CC performance during AVF and TLF.

**Table 3.** Participants’ feedback, and their measured compression rate and depth

| Feedback | CC variables | AVF-led CC, mean (SD) | TLF-led CC, mean (SD) | *p* value<sup>a</sup> |
| --- | --- | --- | --- | --- |
| **Participants' perception** |  |  |  |  |
| Perceived they performed better with TLF (n=18) | Rate, min<sup>-1</sup> | 127.1 (21.4) | 118.7 (15.6) | 0.002 |
|  | Depth, cm | 4.1 (0.8) | 4.1 (0.9) | 0.295 |
| Perceived they performed better with AVF (n=34) | Rate, min<sup>-1</sup> | 118.8 (12.1) | 118.7 (9.8) | 0.936 |
|  | Depth, cm | 4.5 (0.8) | 4.3 (0.9) | 0.228 |
| Perceived they performed well with both methods (n=18) | Rate, min<sup>-1</sup> | 114.5 (10.5) | 113.7 (12.0) | 0.773 |
|  | Depth, cm | 4.7 (0.5) | 4.9 (0.7) | 0.163 |
| **Participants' preference** |  |  |  |  |
| Preferred TLF (n=50) | Rate, min<sup>-1</sup> | 124.1 (19.4) | 118.2 (14.9) | 0.004 |
|  | Depth, cm | 4.3 (0.8) | 4.2 (0.9) | 0.584 |
| Preferred AVF feedback (n=8) | Rate, min<sup>-1</sup> | 121.2 (8.5) | 119.7 (6.5) | 0.569 |
|  | Depth, cm | 4.3 (1.0) | 3.9 (0.8) | 0.007 |
| Did not mind either method (n=12) | Rate, min<sup>-1</sup> | 112.1 (10.1) | 112.8 (9.6) | 0.653 |
|  | Depth, cm | 4.9 (0.6) | 5.1 (0.8) | 0.368 |

<sup>a</sup>Paired t-test.
TLF: team-leader feedback, AVF: audio-visual feedback, CC: chest compression, SD: standard deviation

## Discussion

The 2015 AHA guideline states that high quality CC requires the following: (1) optimal hand position, (2) compressing the lower part of the sternum by at least one-third of the anterior-posterior diameter of the chest (equivalent to 4 cm in infants and 5 cm in adolescents), (3) achieving compression rate of 100 to 120 min^-1^, and (4) allowing for complete chest recoil between each CC. In this study, we focused on the CC rates and depth as these parameters have shown a more significant role in affecting clinical outcome (24). Overall, our study found that the participants’ CC performance with AVF was similar to that with TLF. Although the average CC rate with AVF in our study was statistically significantly higher than that with TLF (121.8 min^-1^ vs. 117.4 min^-1^, *p* = 0.005) and slightly above the recommended target range, this may not be considered significant in real clinical settings.

The proportion of participants performing CPR at CC rates within the recommended target range of 100 to 120 min-1 was similar between AVF and TLF (48.6% and 51.4%, p = 0.824). However, the mean CC depth with both TLF and AVF in this study were below the recommended range, while the mean CC rate inclined toward 120 min^-1^. This corroborated a previous study reporting significant decrease in CC depth as the CC rate increases (25).

With AVF, suboptimal compression depth can be due to difficulty in following the audio-visual prompts that demand competent eyes-ears-hands coordination. The heavy cognitive load resulted in reduced attention capacity towards multiple stimuli during CPR (26). Therefore, despite the AVF indicating inadequate compression depth, participants were inclined to ignore them and continued performing CC without corrective actions. The cognitive loads in CC are intrinsic, germane, and extraneous. Chest compressions are the intrinsic load, whereas the germane load is imposed while using an unfamiliar AVF device. Extraneous load occurs due to the requirement for participants to simultaneously look at the monitor, while listening to voice prompts and fine-tuning their CCs (27, 28). A similar conclusion was made by Brown et al in their study on measuring the task of performing CPR with AVF devices based on the National Aeronautics and Space Administration (NASA) Task Load Index. They reported significantly higher physical burden to CPR providers in multitasking feedback interpretation and formulating corrective measures to improve their compressions (29).

On the other hand, TLF would reduce the extraneous load as participants need to only listen to voice prompts. The germane load would depend on whether participants are familiar to receiving real-time feedback during CC. Team leaders were able to gauge and provide appropriate feedback on the correct compression rate through subjective visual assessment. This suggests that team leaders had conceptual and habitual tacit knowledge of the appropriate CC rate. Tacit knowledge is the implicit knowledge that one possesses based on personal experience (30). It is personal, intuitive, and difficult to be coded, transferred, or taught (31, 32). Schemata on how tacit knowledge and habitual practices influence the management of resuscitation in the ED and other departments have been provided in previous studies (33, 34). Interestingly, assessment and feedback on the CC depth by team leaders in this study were not as accurate as that on CC rate. This may be due to the misidentification of CC depth as adequate at higher compression rates (35).

A recent study (20) demonstrated better CPR quality with feedback device compared to human instructor feedback. Their study method had measured CPR quality as a composite score including correct hand position, adequate depth, compression rate and complete chest recoil. However, similar to our findings, the average CC rate in their study was comparable between feedback device and human instructor feedback, and their human instructor feedback group showed more compliance to CPR guidelines for CC depth.

CPR feedback devices were invented and innovated to automate conventionally human-led resuscitations. In a publication regarding automation of tasks with machines, multiple bottlenecks were identified impeding advancement towards task automation. These bottlenecks involved tasks that require complex manipulation and perception, creative-intelligence tasks, and social-intelligence tasks (36). Leading a resuscitation during CPR in a cardiac arrest is recognized as a complex and highly demanding task requiring case-by-case analysis and insight. Extrapolating these bottlenecks into our study may demonstrate AVF limitations in its ability to only provide objective perception in compression depth and rate. Thus, in more complex cardiac arrest cases, team leaders may be more proficient in employing both vertical and lateral thinking to mitigate suboptimal CC. Lateral thinking is defined as reasoning using an indirect and creative approach that may not be immediately obvious whereas vertical thinking is a thinking that proceeds in a stepwise manner while applying specific rules to reach a goal (37, 38). For instance, in one participant, the team leader had observed and gave feedback of her suboptimal CC attributed by not straightening and locking the elbows.

Furthermore, in our setting, the English language is not a native language and is the second spoken language for most residents. This discrepancy of language proficiency may have resulted in misinterpretation of some AVF prompts. We have noted during the study that most participants were inclined to push faster when the defibrillator audio feedback prompted “push harder”. In contrast, the language conversed by the team leader were a fusion of the native Malay language and conversational English language. Our participants felt that the team leader’s tonal voice was more reassuring as it instilled a sense of confidence and was easier to be understood compared to the machine’s monotonous audio prompts. Effective communication is expressed via spoken words, tone, resonance, pitch modulation and other forms of non-verbal communication (39), some of which, are absent in the AVF method. Perhaps, the socio-cultural variations of vocal intonation in these machines (such as using the local Malay language and dialect) should be considered by the manufacturers to strengthen participants engagement and comprehension.

Previous studies comparing device-led with human-led feedback reported conflicting findings (20, 21). As with Pavo et al, our findings found both methods to be comparable (21). We investigated whether participant’s preference of feedback methods influenced their CC quality, as this has not been previously explored. In our study, participants who perceived that they performed better with TLF and those who preferred TLF performed CC within the recommended range with TLF compared to AVF. However, this was not observed in participants who preferred AVF. Participants who perceived that they performed well with both AVF and TLF, and did not mind one method over the other performed the best.

### Limitations

Our study had some limitations. Firstly, this was a single-center study with participants comprising of ED healthcare personnel and medical students. This may have resulted in selection bias and may not necessarily reflect the overall healthcare providers’ competency in CPR. Although all participants had prior training in BLS, their experience in CPR was likely diverse based on their profession. Secondly, team leaders were not randomized and were allocated to participants based on convenience due to their work schedule. Team leaders also varied in terms of experience and leadership positions. These factors may have resulted in the inter-team leader variability. Thirdly, this study was conducted in a manikin-based simulation setting. This allowed us to standardize the assessment, but it could only represent a real patient scenario to a limited extent. We also chose a shorter duration of CC (i.e. 1 min instead of the 2-min cycle periods in CPR guidelines) to minimize rescuers’ fatigue as our study aimed to assess whether human feedback or feedback device resulted in better CC performance. Results from previous studies showed that CPR quality started to decline after 1 minute due to fatigue (40, 41) regardless of rescuer strength (42), gender, weight, height, or rescuer’s profession (43). Fourthly, we did not have a control group (i.e. CC without feedback). Therefore, we do not know to what extent the CC performance was the effect of TLF or AVF alone or of the participants’ own knowledge and skills. Lastly, a potential Hawthorne effect may have influenced our results as participants were aware that they were being monitored throughout the simulated CC performance.

## Conclusions

In conclusion, our study found similar CC performance between human TLF and machine AVF, and that CC performance may be associated with the method of feedback depending on the provider’s preference. Although AVF provided objective feedback, the need for eyes-ears-hands coordination was perceived as a multitasking challenge for CPR providers to focus on the CC delivery. On the other hand, TLF had a humanistic voice, which was perceived as more reassuring, motivating and easier to follow by CPR providers. The CC performance in our study suggests that more training is needed to improve the quality of CPR regardless of the feedback method used. We suggest that a well-trained team leader could be as effective as an AVF device in leading a good quality CPR.

## Data Availability

The data underlying the results presented in the study are available from the corresponding author via email at

## Acknowledgements

The management and medical staff of the Emergency Department of the study center.

